# Increasing transmission of dengue virus across ecologically diverse regions of Ecuador and associated risk factors

**DOI:** 10.1101/2023.05.25.23290519

**Authors:** Leah C. Katzelnick, Emmanuelle Quentin, Savannah Colston, Thien-An Ha, Paulina Andrade, Joseph N.S. Eisenberg, Patricio Ponce, Josefina Coloma, Varsovia Cevallos

**Author notes:** Corresponding authors: Leah Katzelnick, Josefina Coloma, and Varsovia Cevallos.

## Abstract

The distribution and intensity of viral diseases transmitted by *Aedes aegypti* mosquitoes, including dengue, have rapidly increased over the last century. Ecuador is an interesting country to study drivers of dengue virus (DENV) transmission given it has multiple ecologically and demographically distinct regions. Here, we analyze province-level age-stratified dengue prevalence data from 2000-2019 using catalytic models to estimate the force of infection of DENV over eight decades and across provinces in Ecuador. We found that provinces established endemic DENV transmission at different time periods. Coastal provinces with the largest and most connected cities had the earliest and highest increase in DENV transmission, starting around 1980 and continuing to the present. In contrast, remote and rural areas with reduced access, like the northern coast and the Amazon regions, experienced a rise in DENV transmission and endemicity only in the last 10 to 20 years. The newly introduced chikungunya and Zika viruses have distinct age-specific prevalence distributions consistent with recent emergence across all provinces. We evaluated factors to the resolution of 1 hectare associated with geographic differences in vector suitability and arbovirus disease in the last 10 years by modeling 11,693 *A aegypti* presence points and 73,550 arbovirus cases. In total, 56% of the population of Ecuador lives in areas with high risk of *Aedes aegypti*. Most suitable provinces had hotspots for arbovirus disease risk, with population size, elevation, sewage connection, trash collection, and access to water as important determinants. Our investigation serves as a case study of the changes driving the expansion of DENV and other arboviruses globally and suggest that control efforts should be expanded to semi-urban and rural areas and to historically isolated regions to counteract increasing dengue outbreaks.

**AUTHOR SUMMARY:** The factors driving the increasing burden of arboviruses like dengue virus are not fully understood. In this study, we measured changes in dengue virus transmission intensity and arbovirus disease risk across Ecuador, an ecologically and demographically diverse South American country. We found that differences in the distribution of dengue cases could be explained by changes in transmission of dengue virus over time: transmission was limited to coastal provinces with large cities between 1980-2000, expanding thereafter to higher elevation areas and ecologically suitable but previously geographically and socially isolated provinces. We also used species and disease distribution mapping to show that both urban and rural areas in Ecuador are at medium to high risk for *Aedes aegypti* presence and arbovirus disease risk, with population size, precipitation, elevation, sewage connection, trash removal, and access to water as strong predictors. Our investigation reveals changes driving the expansion of dengue and other arboviruses globally and provides an approach for identifying areas at early stages of establishing endemic transmission that should be targeted for intense preventative efforts to avert future epidemics.

## INTRODUCTION

Arboviruses transmitted by the *Aedes aegypti* mosquito (Culicidae), including the flaviviruses yellow fever virus, dengue viruses 1-4 (DENV1-4), and Zika virus (ZIKV) and alphavirus chikungunya virus (CHIKV), infect millions of people globally each year and cause a spectrum of life-threatening diseases with long-term sequelae, including hemorrhagic fevers, arthritis, and severe congenital abnormalities [1, 2]. In the 19^th^ and 20^th^ centuries, explosive epidemics of arboviral disease primarily affected large population centers in tropical regions but in the 20^th^ into the 21^st^ century, arboviruses transmission worsened in previously affected areas and expanded into new regions [3–5]. As a result, dengue is one of the few tropical diseases with an increasing global burden over the last 20 years [6]. The distribution of other arboviruses has also increased over the last ten years as ZIKV and CHIKV were introduced to the Americas and caused back-to-back continent-wide pandemics [7, 8].

Tracking factors driving the global expansion of arboviruses like DENV is limited by the quality of disease surveillance. In many regions, dengue prevalence remains low but whether this is attributable to low reporting rates as opposed to low disease burden is difficult to determine. Recent and historical work suggests that the age distribution of incident cases is a more accurate measure of transmission intensity and is more robust to common variations in surveillance quality than aggregate case count data [9–11]. Specifically, the force of infection (FoI), defined as the per-capita rate at which susceptible individuals become infected in a population within a given period, can be directly estimated from age-stratified prevalence data and provides a useful measure of transmission intensity. Vector surveillance combined with statistical prediction methods of climate and other factors can also serve as an early warning system for human dengue outbreaks [12, 13].

The ecological and demographic drivers of arboviral disease can be studied within a single diverse country like Ecuador, a South American country with multiple distinct regions capable of sustained arbovirus transmission. Both the tropical coastal lowlands to the east and the Amazon to the west have suitable conditions for *Ades aegypti* but are divided geographically by the Andean mountains and have limited transportation connectivity. Further, each region is bisected by the equator, resulting in distinct seasonality between north and south and thus offset arbovirus transmission periods.

Ecuador has a long history of arbovirus transmission, with the first case of yellow fever recorded in 1740 and the first outbreak in 1842 [14, 15]. Between 1946 and 1970, eradication campaigns targeted the vectors for yellow fever and malaria, resulting in a major reduction in *Aedes* populations. *Aedes aegypti* soon returned, followed by the re-introduction of dengue [3]. National dengue outbreaks occurred in Ecuador in 1996, 2000, 2005, 2010, 2012, and 2014 and since 2000, the four DENV serotypes have been detected in all regions [16, 17]. A recent phylogenetic analysis showed both recent and historical introductions of DENV1 and DENV2 in Ecuador that originated from Venezuela and Colombia, while chikungunya was introduced through the Caribbean [18, 19]

Here, we modeled province-level age-stratified dengue prevalence data from 2000-2019 to estimate changes in the DENV transmission intensity for nearly 80 years and across provinces in Ecuador. We then evaluated factors associated with geographic differences in vector suitability and arbovirus disease in the last ten years by modeling 11,693 *Aedes aegypti* presence points and 73,550 arbovirus cases. We identify provinces at different stages of establishing endemic DENV transmission as well as hotspots for vector and disease risk.

## MATERIALS AND METHODS

### Ethics statement

Human subjects data sets were from anonymized public health surveillance data collected by the Ecuadorian Ministry of Health as part of routine care. The Instituto Nacional de Estadística y Censos (INEC) data used in this study is freely available from an institutional website (https://www.ecuadorencifras.gob.ec/camas-y-egresos-hospitalarios/) and is completely anonymous. According to the international good clinical and research practice and in accordance with Ecuadorian legislation on clinical investigation, ethical approval by an institutional review board was not required. The public surveillance data collected by the Ecuador Ministry of Health (Sistema Nacional de Vigilancia en Salud Pública Ecuador, ViEpi data) was made available for research purposes without confidential information (anonymous), under the “Organic Law of Transparency and Access to Public Information” (Article 4 of Ley Orgánica de Transparencia y Acceso a la Información Pública, LOTAIP, Law 24 of Article 81 of the Ecuadorian Constitution).

### Temperature and precipitation data

Annual province-level temperature and precipitation data from 2000-2019 were derived from the Universidad Tecnológica Equinoccial (UTE)/Centro de Investigación en Salud Pública y Epidemiología Clínica (CISPEC) database. Trend lines were fitted using local weighted regression (LOESS) models. Province level monthly arbovirus case distributions are for dengue, Zika, and chikungunya cases from 2014 and 2017 in the ViEpi database (n=41,777 total cases, described below).

### Age-stratified dengue prevalence data and catalytic models

Dengue prevalence was estimated from national surveillance data collected by INEC. The INEC dataset included all discharge data for hospitalized cases classified as vector-borne viral diseases and/or hemorrhagic fevers by International Statistical Classification of Diseases (ICD). Data were available from 2000 to 2019 and were grouped by age, sex, and province. For analysis, ICD codes A90 (Classic Dengue Fever) and A91 (Dengue Hemorrhagic Fever) were grouped as dengue cases (1997 WHO criteria for severe dengue disease). Ages were binned into 18 age groups (<1, 1-4, 5-9, 10-14, 15-19, 20-24, 25-29, 30-34, 35-39, 40-44, 45-49, 50-54, 54-59, 60-64, 65-69, 70-74, 75-78, and ≥80 years of age) for analysis.

To estimate prevalence, we used provincial demographic data as our denominator. National census data from 1991, 2000, and 2010 were available for each province and age group; we used interpolation to estimate the age-specific population size for each year from 2000 to 2019. There were sufficient case counts in all provinces in the coastal region (Esmeraldas, Manabí, Guayas/Santa Elena, El Oro, Pichincha/Santo Domingo de los Tsáchilas, Los Ríos) to enable province-level DENV FoI estimation. Although Santa Elena was separated from Guayas province and Santo Domingo de los Tsáchilas was separated from the Pichincha province in 2007, we leave them as combined provinces in our analyses to maintain consistency across years. Due to lower absolute case counts, we combined the case data for all Amazon provinces: Sucumbíos, Napo, Orellana, Pastaza, Morona Santiago, and Zamora Chinchipe. To evaluate the relative contribution of each age group to the total yearly prevalence, we plotted the age-stratified dengue prevalence normalized by the total prevalence in each year to account for differences in absolute epidemic magnitude. Data were grouped into two periods (2000-2009 and 2010-2019) to visualize temporal changes in transmission. To compare distributions, data were fit with a LOESS curve with 95% confidence intervals. We used the age-adjusted prevalence data to estimate medians and interquartile range of ages by province for each 10-year period.

We used catalytic models to estimate historical and current FoI of DENV for each province based on age-stratified prevalence using the method described by Rodriguez-Barraquer et al. [9], as follows.

The fraction susceptible to all four DENV serotypes at age *a* and time *t* is:

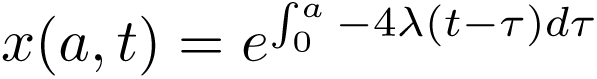

Where, λ(*t*) is the average serotype-specific FoI and τ is the time since birth. The fraction that has been infected with one serotype and remain susceptible to the other four serotypes (monotypic immune) is:

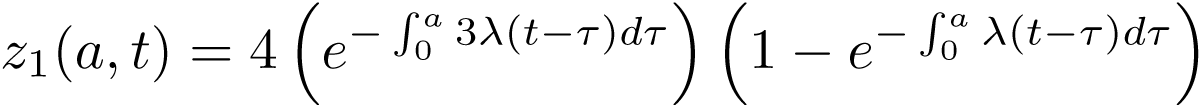

Consistent with prior analyses, we assume that the age-specific prevalence of dengue is driven by those experiencing their second DENV infection [20–22]. The at-risk population thus consists of individuals with primary DENV immunity, i.e. individuals infected with one DENV serotype but still at risk of infection with any of the other three serotypes. Thus, the age-specific prevalence for dengue is assumed to reflect the monotypic group experiencing their second infection with any of the other serotypes:

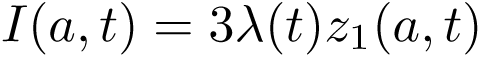

The expected number of reported cases at age *a* and time *t*, Λ(*a*, *t*), is a function of prevalence, as well as the population size *P(a,t*) and reporting rate φ(*t*):

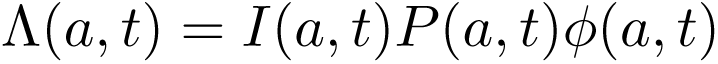

We assume the observed number of reported cases at age *a* and time *t*, *C(a,t)*, follows a Poisson distribution with the likelihood:

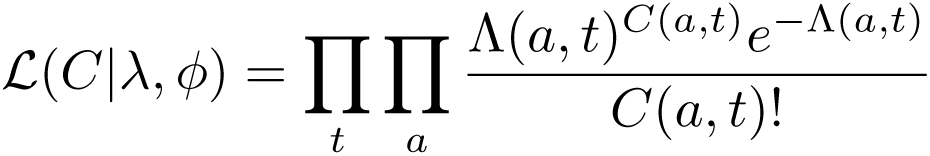

Models were fit with Rstan [23] using the Bayesian Markov Chain Monte Carlo framework in R [24]. We ran four independent Markov chains with 10,000 iterations of warmup followed by 20,000 iterations for estimating parameters. Parameters were evaluated for convergence using the split *^R^^* statistic (values of 1 were assumed to indicate convergence) as well as visual inspection.

All FoI estimates are shown as means with 95% credible intervals. The original authors of this code [9] note that given the large numbers for prevalence, credible intervals may be systematically underestimated.

### Comparison of dengue, chikungunya, and Zika age-stratified prevalence

We measured differences in arbovirus age-stratified disease prevalence using the ViEpi dataset. The ViEpi dataset included de-identified individual case information for 2014 (only November to December), 2015 (full year), 2016 (full year), and 2017 (only January to April) meeting the following case definitions: (1) Dengue with Warning Signs or Severe Dengue, (2) chikungunya, and (3) Zika. The dataset included the following information for each case: age, sex, nationality, home district, province, canton, parish, date of illness, whether and how the case was confirmed (laboratory confirmation, clinical, contact case), hospitalization status, and outcome of case. Cases were grouped by the same age categories as indicated above for the INEC dataset. We use suspected, and not just laboratory-confirmed cases, as there is an age-specific bias in the cases that were confirmed. Notably, the prevalence of chikungunya and Zika cases in infants and for Zika, reproductive age individuals, is higher than would be expected, possibly due to greater confirmation in these age groups.

For our didactic example of CHIKV and ZIKV FoI, we assumed the age-specific prevalence reflects the fully susceptible group experiencing a first infection:

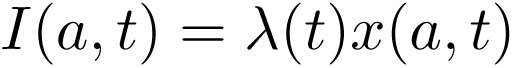

### Entomological and disease data

*Aedes aegypti* presence was measured as part of the INSPI-SENESCYT Project “National Vigilance and Early Warning System for Vector Control of Dengue-Yellow Fever” (SATVEC PIC-12-INH-002). *Aedes aegypti* mosquito samples were collected from 2013 to 2018, at 1785 collection sites in Ecuador. In total 946 records correspond to collections during the rainy season and 839 during the dry season. Coordinates for all collection records were obtained. Samples were collected and transported to the laboratory for specimen identification. For arbovirus disease mapping, dengue and chikungunya cases reported in the Ministry of Public Health surveillance data in 2015 in the ViEpi dataset were used as a measure of disease presence across Ecuador.

### Species and dengue case distribution mapping using maximum entropy models

We used the confirmed vector and disease presence data in maximum entropy models to evaluate the species distributions for *Aedes aegypti* and dengue and Chikungunya across Ecuador using the MaxEnt software (version 3.3.3), available at: https://biodiversityinformatics.amnh.org/open_source/maxent/ [25, 26]. The maximum entropy model uses environmental and social layers to estimate the probability of presence of a disease or species within each raster unit to predict the distribution of a disease or species across the map. MaxEnt was selected because it is widely used, has been proven to be one of the best models for species distributions, and has a GIS interface within the TerrSet GIS software (Clark Labs). We used a similar method to that described previously for characterizing the distribution of *Aedes Albopictus* in Ecuador [27]. We trained the model on 75% of presence points, with 25% for validation, and 10 bootstrap repetitions. Model goodness-of-fit was estimated as Area Under the Curve (AUC). Variables previously identified as predictors of arbovirus transmission were included as layers in the model: two physiographic variables (elevation, slope), eight demographic and social variables derived from available census data (literacy, employment, overcrowding, population density, housing condition, water access, sewerage, garbage collection), and three environmental/climatic variables (precipitation, Daytime Land Surface Temperature, and the Enhanced Vegetation Index). All variables were downscaled at a resolution of 1 hectare: regional data disaggregation was based on the density population map for arboviral case points and a uniform indicator at the census tract level for the INEC census database. The input environmental geodatabase was obtained by processing EarthData images (NASA: https://www.earthdata.nasa.gov/), with the spatiotemporal series of monthly images from 2010 to 2018 imported and downscaled to Ecuador at 1 hectare resolution. As the series of environmental variables was too large to include in the MaxEnt model, we applied an orthogonal transformation of n-dimensional image data using principal component analysis (PCA) to produces a new set of images (components) that are uncorrelated with one another and ordered with respect to the amount of variation (information) they represent from the original image set.

These techniques were implemented in TerrSet. As we were working with a cube X-Y-T, the PCA could be adapted to the various dimensions. For the three climatic parameters (temperature, precipitation, and vegetation) we used two PCA components in T mode and 2 PCA loadings in S-mode. Using this approach, the model was not affected by layers that would not be explicative; indeed, it can occur that a variable is useful in the coast and not in the highlands, a benefit of working spatially.

## RESULTS

### Geography, climate, and demographic characteristics of Ecuador

Ecuador is a country in South America of 283,561 square kilometers and a total population size of just over 17 million people. Ecuador is divided down the center by the Andes mountains; provinces in this region are at an altitude too high to support *Aedes* populations and DENV transmission (**Fig. 1**). To the west of the Andes are the coastal lowlands, which border the Pacific Ocean, are densely populated, have high temperatures and humidity, and distinct wet and dry seasons. The provinces of Esmeraldas, Manabí, Santa Elena/Guayas (we combine these two provinces in our analysis), Los Ríos and El Oro consist primarily of lowland regions while Santo Domingo de los Tsáchilas/Pichincha (we combine these two provinces in our analysis) contain both lower and higher elevation areas. Esmeraldas is the only province north of the equator, and both Esmeraldas and higher-elevation Tsáchilas/Pichincha have lower and more stable year-round temperatures than other provinces in this region (**Fig. 1**). In all coastal provinces, the rainy season extends from November through May, with the highest number of arboviral disease cases occurring between March and July. Six of the eight cities in Ecuador with populations larger than 200,000 are on the coast, including the major port cities Guayaquil (Guayas) and Manta (Manabí). The six eastern provinces of Ecuador are part of the Amazon, with a tropical climate but lower population density and no cities with more than 120,000 people. Precipitation is high year-round, with slightly more rainfall from March to July and in the northern compared to the southern Amazonian provinces. Consistently, there is less seasonality of arbovirus transmission in the Amazon than the coastal provinces. The Galapagos Islands constitute the fourth region of Ecuador and while cases are sporadically reported, there is little evidence of sustained transmission.

**Fig. 1.**
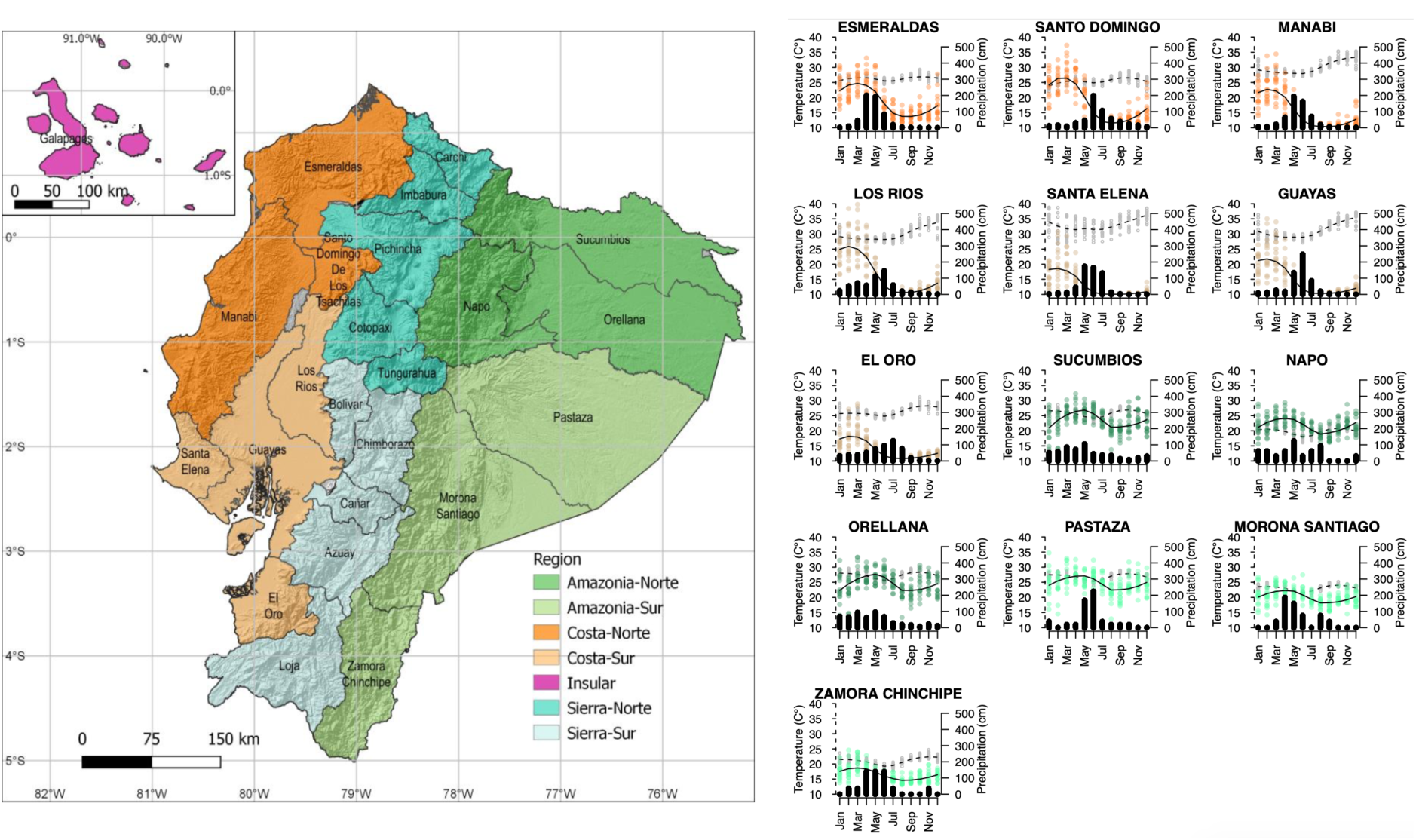
Geography and arbovirus seasonality of Ecuador. **(Left)** Map of Ecuador, colored to indicate geographic and climate regions (north vs. south; coast, mountains, vs. Amazon). **(Right)** Temperature (grey dots and dotted lines), precipitation (dots colored by region, solid black line), and monthly distribution of arbovirus cases (black histograms). Temperature and precipitation data are shown for 2005-2019, while monthly arbovirus case distributions are dengue, Zika, and chikungunya cases from 2014 and 2017 in the ViEpi dataset.

### Age-distributions of dengue prevalence vary by province and over time

We first measured temporal and geographic differences in DENV transmission intensity by estimating age-stratified dengue prevalence for each province between 2000-2019. In general, the age-distribution of disease reflects the average age at which individuals experience their first infection. However, because primary DENV infections are less likely to cause symptoms or disease severe enough to elicit a hospital visit, most cases captured by surveillance represent secondary DENV infections. We estimated dengue prevalence using national surveillance of Dengue Fever and Dengue Hemorrhagic Fever cases and interpolated age-specific province-level population size data. For data visualization, data were grouped into two 10-year periods, 2000-2009 and 2010-2019 (**Fig. 2**).

**Fig. 2.**
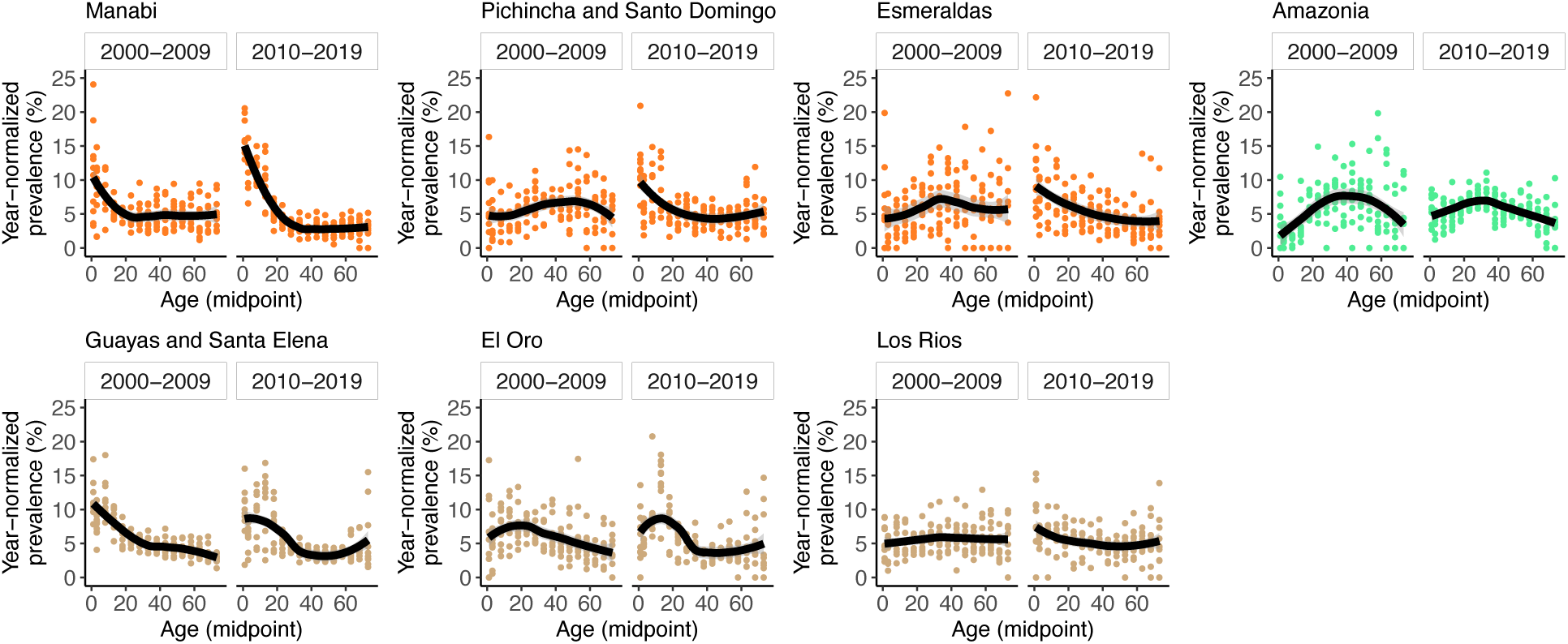
Age-specific prevalence of dengue, normalized by year, for each province and period. Data are fitted using LOESS regression with 95% confidence intervals. Data are from the INEC database.

The median age of the population in each province ranged from 20-26 throughout the 20-year period, with the lowest median age in the Amazon and highest age in Guayas/Santa Elena and Pichincha/Santo Domingo (**Table 1**). However, the first decade of data had different features compared to the second decade of data. For example, from 2000 to 2009, the population-dense coastal provinces of Guayas/Santa Elena, El Oro, and Manabí had the lowest median age of dengue cases (28-36 years) whereas in provinces with higher elevation or that were more rural and inaccessible (Esmeraldas and the Amazon), the median dengue prevalence occurred at older ages (41-43 years). All provinces except Guayas/Santa Elena experienced a drop in median dengue prevalence between 2000-2009 and 2010-2019, with the largest change in Manabí (by 17 years), as well as large changes in Esmeraldas, the Amazon, and Pichincha/Santo Domingo (6-9 years).

**Table 1.** Median and interquartile range (IQR) of population-adjusted dengue prevalence by province, from 2000-2009 and 2010-2019. Median and IQR of the population during the same period are shown for reference.

| Province | Prevalence: median<br>(IQR) |  | Population: median<br>(IQR) |  |
| --- | --- | --- | --- | --- |
|  | 2000-2009 | 2010-2019 | 2000-2009 | 2010-2019 |
| Manabí | 36 (13-58) | 19 (8-48) | 24 (12-41) | 25 (12-43) |
| Pichincha/Santo Domingo | 43 (25-60) | 35 (14-59) | 26 (13-42) | 26 (13-42) |
| Esmeraldas | 41 (25-59) | 32 (14-56) | 22 (11-39) | 22 (11-40) |
| Amazonia | 43 (27-58) | 37 (22-57) | 20 (9-36) | 21 (10-37) |
| Guayas/Santa Elena | 28 (11-51) | 28 (13-58) | 26 (13-42) | 26 (12-43) |
| El Oro | 32 (16-51) | 28 (13-57) | 25 (12-42) | 26 (13-44) |
| Los Ríos | 41 (22-60) | 39 (18-62) | 24 (11-41) | 25 (12-42) |

### Models of age-stratified dengue prevalence reveal historical and geographic changes in DENV transmission

We assumed that the age distributions of dengue prevalence by province were due to both current and historical transmission of DENV in each region. We estimated the FoI of DENV for 10-year increments back to 1941, covering the earliest period when individuals in the dataset were alive. We modeled annual age-stratified dengue prevalence directly using Bayesian models based on the method described by Rodriguez-Barraquer et al. [9]. We assumed the surveillance data reflect prevalence of second DENV infections, as has been assumed previously [20–22]. We observed that all provinces and regions had minimal DENV transmission for the period of 1941 to 1979 (**Fig. 3**). Consistent with this observation, from 1946 to 1970, continent-wide *Aedes* eradication campaigns were underway. A program was implemented in 1958 to specifically monitor *Aedes aegypti* [3]. No dengue cases were recorded in Ecuador during this time, but *Aedes* successfully reinfested the cities in Guayas and Manabí between 1977 and 1985. Our FoI models estimated that between 1980 to 1989, DENV transmission intensity began to rise in Guayas/Santa Elena and El Oro (**Fig. 3**). Then from 1990-1999, high DENV transmission was observed in Guayas/Santa Elena, El Oro, and Manabí. These results are consistent with the fact that dengue was first documented in Ecuador in 1985, and major outbreaks were observed in Guayas in 1988, 1990, and 1993. At this time, dengue was re-emerging across the Americas, and the introduction of multiple serotypes resulted in the first observed cases of Dengue Hemorrhagic Fever/Dengue Shock Syndrome [3, 4, 28]. The largest and earliest dengue outbreaks were concentrated in the large coastal cities in Guayas and Manabí, consistent with our estimated timeline for the re-introduction of dengue to Ecuador.

**Fig. 3.**
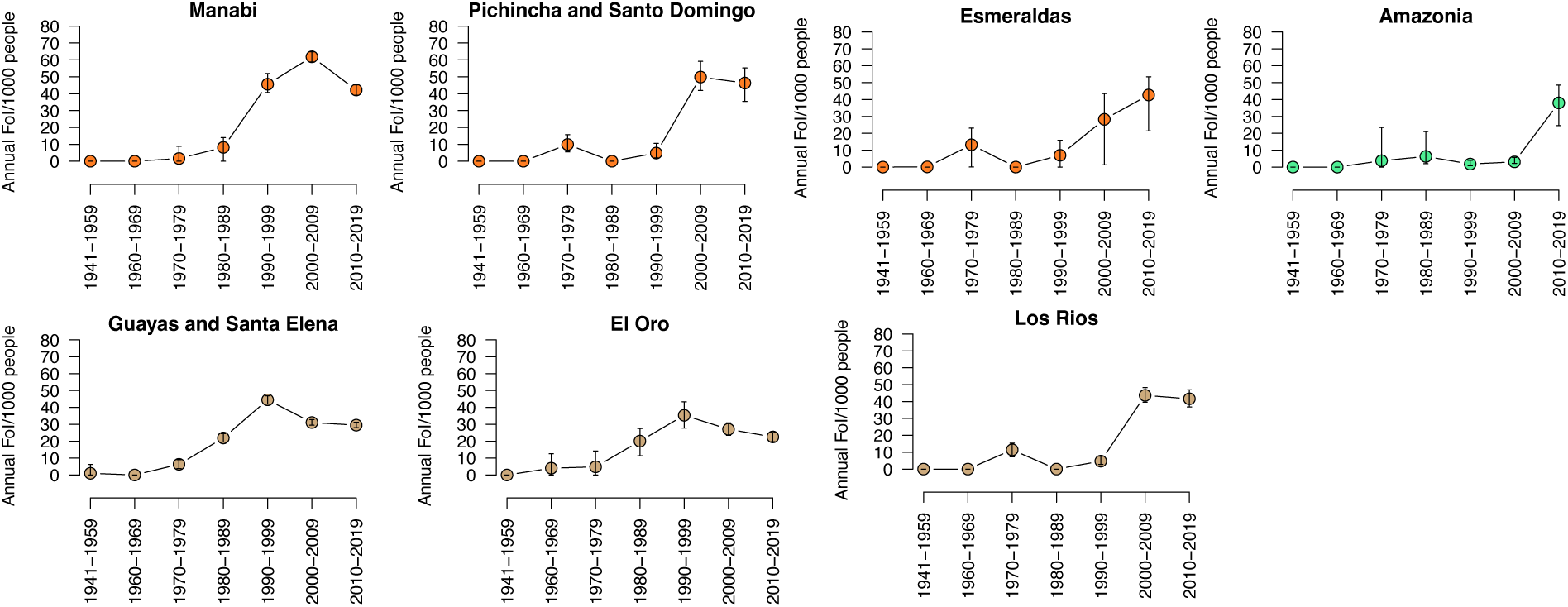
DENV FoI estimates for provinces across Ecuador. Point estimates are colored by geographic region, 95% credible intervals are shown.

Other regions with suitable ecological conditions for DENV transmission in Ecuador have not recorded major epidemics. Consistent with this epidemiological data, we found that the early rise of DENV in the southern coastal provinces was not observed in the northern coastal province of Esmeraldas, the higher elevation coastal provinces of Los Ríos and Pichincha/Santo Domingo de los Tsáchilas, or the Amazon provinces, which had very low levels of DENV transmission from 1990-1999 (FoI: ≤10 per 1000 people). These provinces had fewer dense urban settings and were more isolated before the year 2000, with limited road infrastructure. Pichincha/Santo Domingo, Los Ríos, and Esmeraldas had high DENV transmission intensity in 2000-2009 and 2010-2019 (FoI: 40-50 per 1000 people). The Amazon provinces were most delayed, experiencing a rise in DENV transmission only in the last 10 years (40 per 1000 people). Thus, we observe the provinces of Ecuador at different stages of endemicity, with some only recently experiencing increases in transmission intensity.

### Age-distributions of recently emerged arboviral diseases reflect risk across ages

CHIKV was introduced to Ecuador in 2014 and caused a major outbreak in 2015, soon followed by a national Zika epidemic in 2016 [8, 18]. We expected that the age distributions of these new arboviral diseases would be similar across provinces, consistent with their recent introduction. We measured the age and disease-specific prevalence using data from a separate surveillance system, ViEpi, consisting of laboratory-confirmed and suspected cases of chikungunya, Zika, and Dengue with Warning Signs/Severe Dengue observed between 2015 and 2016. Across provinces, Zika and chikungunya prevalence was mid-aged centered, consistent with recent emergence, although in some regions, Zika prevalence was more concentrated in reproductive-age individuals, likely due to a bias in laboratory confirmation of cases (**Fig. 4**). Notably, the age-distributions of chikungunya and Zika prevalence more closely resembled the age-distributions of dengue prevalence in Esmeraldas, Los Ríos, Pichincha/Santo Domingo, and the Amazon, but were starkly different in Guayas/Santa Elena, El Oro, and Manabí. These findings further support the hypothesis that heterogeneity in the age distribution of dengue prevalence across Ecuador reflects historical differences in the time of DENV emergence and the transition to endemicity.

**Fig. 4.**
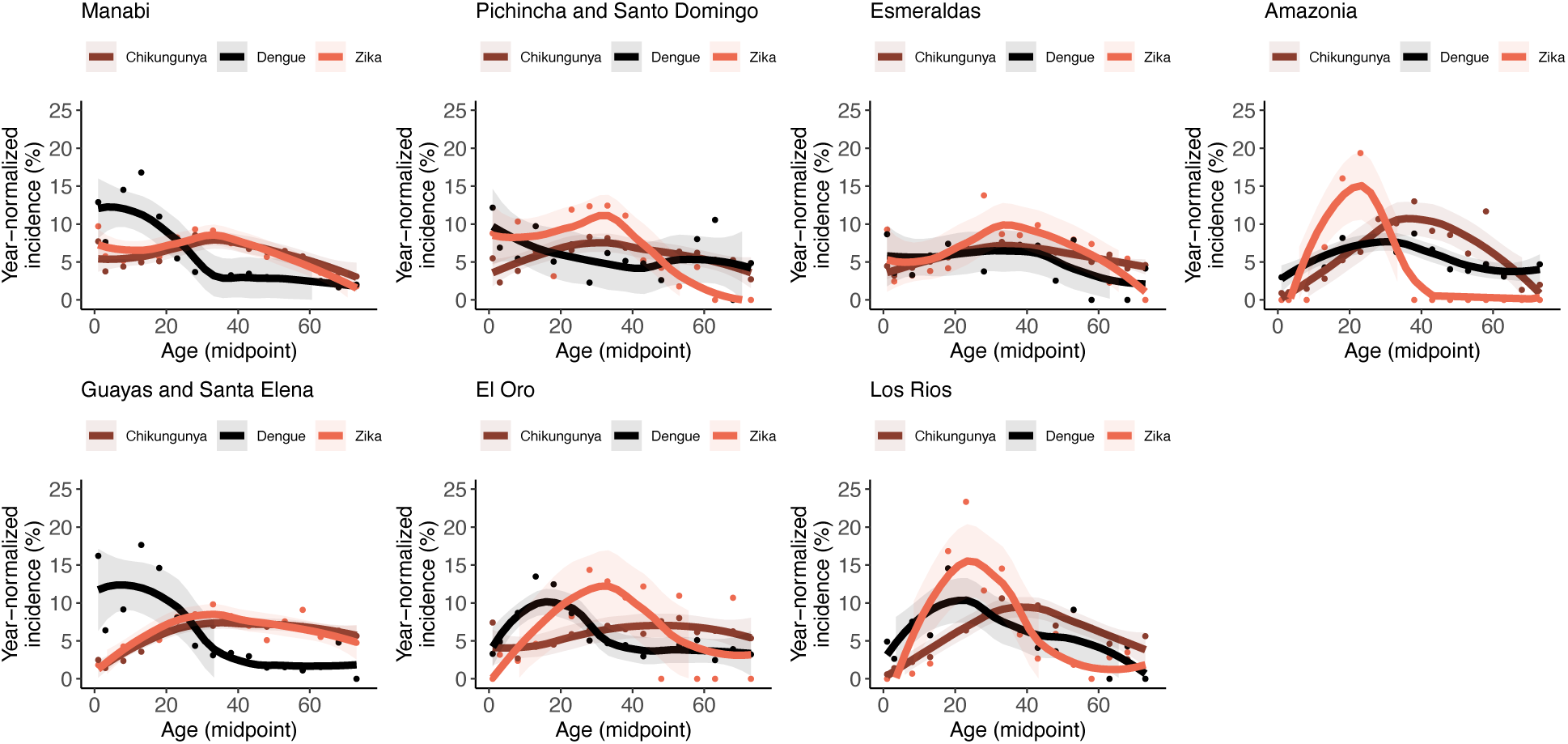
Age-specific prevalence of laboratory-confirmed and suspected Dengue with Warning Signs/Severe Dengue, chikungunya and Zika for each province in 2015-2016. Data are fitted using LOESS regression with 95% confidence intervals. In some provinces, unusual distributions for Zika suggest case-confirmation focused on specific age groups.

It is important to note that the age-specific prevalence distribution for arboviruses such as chikungunya and Zika, where a first infection causes disease, is expected to differ from dengue, where a second infection is most likely to cause disease as illustrated in Figure 5 using a theoretical example (**Fig. 5**). For all three diseases, as years pass since emergence, the age distribution shifts from mid-age distributed to younger ages as older individuals become immune and disease only affects young, naïve individuals. While for chikungunya and Zika, explosive epidemics are observed immediately, for dengue, it takes multiple years before high prevalence is observed across age groups as individuals experience second infections. Further, the shape of the age distribution of cases at endemicity is distinct for dengue compared with chikungunya and Zika, reflecting these differences in the susceptible population.

**Fig. 5.**
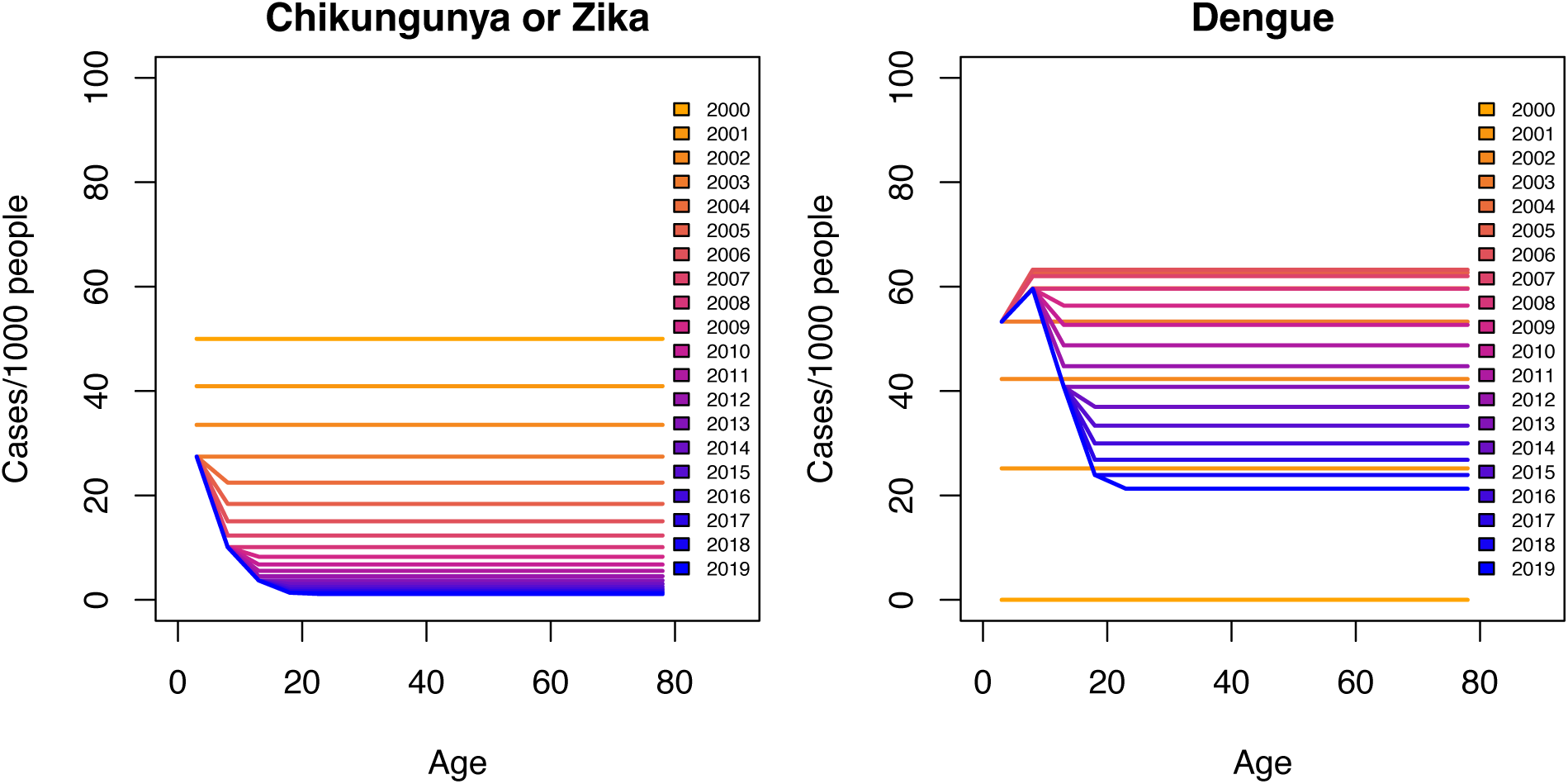
Age-stratified prevalence of different arboviral diseases by time since emergence. Expected changes in age-specific prevalence for a viral disease that emerged 20 years ago with an annual FoI of 50/1000, assuming the first infection causes disease (e.g. like chikungunya or Zika) vs. when there are four circulating viruses that emerged 20 years ago each with an annual FoI of 50/1000 and the second infection causes disease (e.g. like dengue).

### Determinants of *Aedes* and arbovirus risk help explain geographic variation in DENV transmission intensity

To evaluate which environmental, geologic, and demographic factors were associated with the observed geographic differences in DENV transmission intensity, we used maximum entropy models to identify predictors of *Aedes aegypti* and arboviral disease presence data. We sampled immature and adult stages of *Aedes aegypti* mosquitoes in 1785 distinct collection sites, encompassing all 24 provinces in Ecuador, elevation ranging from 0 to 1650 meters above sea level, and during rainy and dry seasons over five years (2013-2018) for a total of n=11,693 distinct samples (**Fig. 6**). The percentage of sites with presence of *Aedes aegypti* in the coastal provinces was higher (35.9%) than in the Amazonia provinces (13.4%) but *Aedes aegypti* was present at higher altitudes in the eastern Amazon basin (1650 meters) than the coastal region (1000 meters). In our maximum entropy models, 55.5% (123,087 km^2^) of the territory was suitable (medium or high risk) for *Aedes aegypti*. Most of the coastal lowlands had medium risk of for *Aedes aegypti*, with high-risk zones in more populated areas. Almost all of Guayas and El Oro were at high risk, while large sections of Manabí, Esmeraldas, Pichincha/Santo Domingo, and Los Ríos were also at high risk. Much of the Amazon was predicted to be at low risk, although the northern Amazon and Amazon regions closer to the Andes were also at medium to high risk for *Aedes aegypti*.

**Fig. 6.**
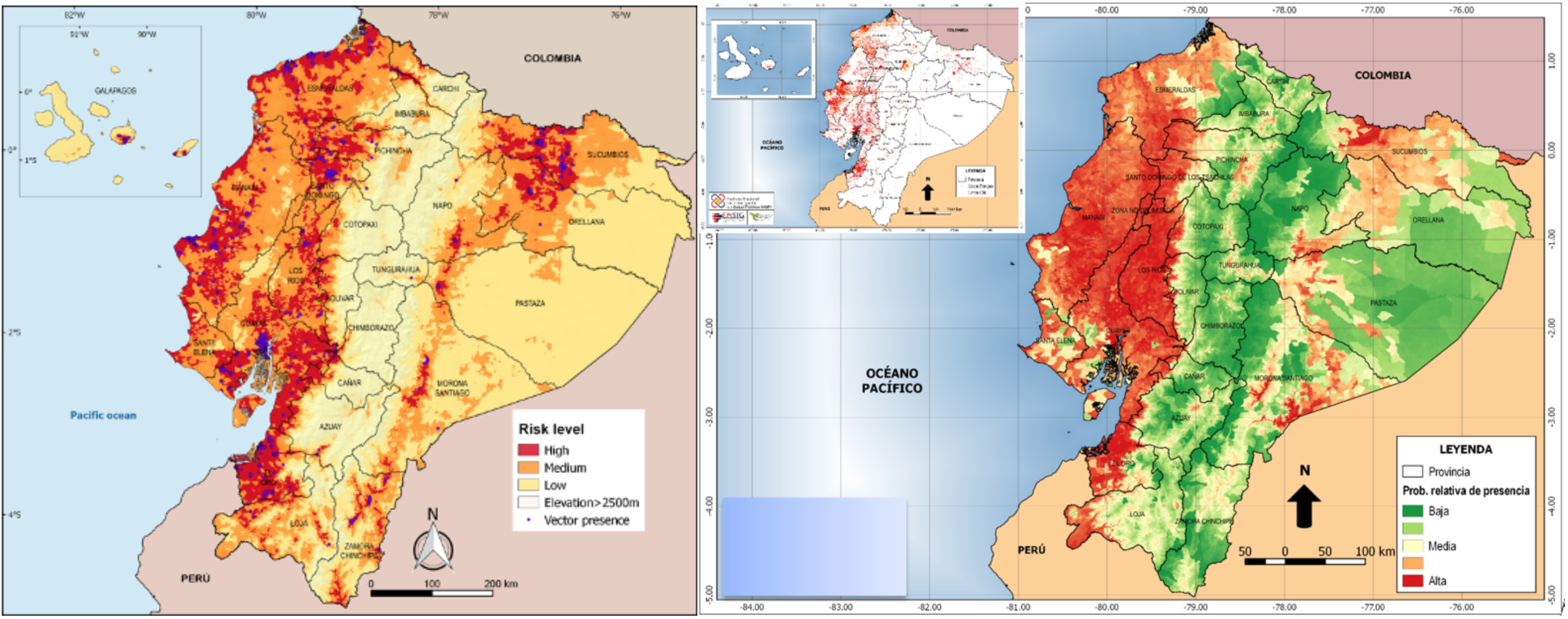
Maximum entropy geographic models using environmental, geologic, and demographic variables to predict *Aedes aegypti* (left) and arboviral disease (right) presence data. Mosquito data: immature and adult stages of *Aedes aegypti* mosquitoes at 1785 distinct collection sites, encompassing all 24 provinces, elevation ranging from 0 to 1650 meters above sea level, and during rainy and dry seasons over five years (2013-2018) for a total of n=11,693 distinct samples. Arbovirus disease prevalence: national dengue and chikungunya cases in 2015 (n=73,550).

For our map of arbovirus disease prevalence, we used dengue and chikungunya cases in 2015 (n=73,550) as a measure of disease presence across Ecuador and maximum entropy models with the same predictor variables (**Fig. 6)**. The regions identified as being suitable for arbovirus disease matched those identified for *Aedes aegypti* risk: most of the coastal region was at high risk for arbovirus transmission, while similar, although not fully overlapping areas across the Amazon were at medium to high risk. Interestingly, an area in the southern Amazon province of Morona Santiago had higher risk for arbovirus transmission risk than predicted by *Aedes aegypti* maps.

The principal variables that predicted *Aedes aegypti* presence also predicted arbovirus disease, including population density (63.3% contribution for *Aedes* risk, 52.8% for arbovirus risk), garbage collection (14.3% and 7.7%), elevation (11.2% and 19.9%) (**Table 2**). However, precipitation was a much stronger factor for arbovirus risk (12.8%) than for *Aedes* risk (1.0%). Both *Aedes* and arbovirus risk were associated to a lesser extent with water access (3.6% and 1.1%) and sewage connection (1.6% and 2.8%), while only *Aedes* was affected by vegetation (1.8%) and house quality (0.7%) and arbovirus disease with house crowding (0.7%).

**Table 2.** Variable importance of risk factors in the *Aedes aegypti* presence data and dengue and chikungunya case presence.

| Aedes model<br>AUC full model: 0.977 |  | Dengue and chikungunya model<br>AUC full model: 0.782 |  |
| --- | --- | --- | --- |
| Variable | Percent contribution | Variable | Percent contribution |
| Population | 63.3 | Population | 52.8 |
| Garbage collection | 14.3 | Elevation | 19.9 |
| Elevation | 11.2 | Precipitation | 12.8 |
| Water access | 3.6 | Garbage collection | 7.7 |
| Vegetation | 1.8 | Sewage connection | 2.8 |
| Sewage connection | 1.6 | Water access | 1.1 |
| Precipitation | 1.0 | House crowding | 0.7 |
| Housing quality | 0.7 | Other variables | 2.2 |
| Other variables | 2.5 |  |  |

## DISCUSSION

In this study, we described changes in the transmission intensity of DENV across ecologically diverse provinces of Ecuador over nearly 80 years and identified multiple environmental, demographic, and social factors associated with the distribution of the vector and arbovirus disease. We found that the highest DENV FoI was observed in the southern coastal provinces over 20-30 years ago, whereas regions that were historically more rural and isolated experienced their highest FoI in the last 10 years. Our geographic models show that the coastal provinces and Amazon have high vector and disease risk areas, and that population density, garbage collection, elevation, and precipitation explain the most variation (>10%) in vector and arbovirus distributions. Our estimates of the FoI of DENV complement and enrich the documented record of DENV emergence across Ecuador and serve as a case study of the changes driving the expansion of arboviruses globally.

We found that DENV transmission in Ecuador increased as dengue resurged across the Americas, first in the southern coastal provinces with major port cities, then to other connected regions in the coastal lowlands. DENV only spread to more isolated regions of Ecuador in the 21^st^ century, possibly driven by increased human movement. Starting near the turn of the 21^st^ century, the government of Ecuador began to heavily invest in roads and infrastructure. The new roads increased connectivity to previously isolated regions, including major cities in Esmeraldas and the Amazon, as well as to more rural areas in all provinces [29, 30]. These changes may have driven the uptick in DENV FoI in Esmeraldas and the Amazon, which were not previously considered high risk areas. Further, higher elevation regions like Pichincha/Santo Domingo and Los Ríos experienced an increase in dengue, possibly due to changes in global temperature expanding the range of *Aedes aegypti* [13]. In total, 55.5% of Ecuador currently has favorable conditions for *Aedes aegypti,* including the provinces we identify here as experiencing increases in DENV transmission intensity in the last 10 years.

As previously low-population rural areas become increasingly urbanized and connected to other rural areas and to major cities, their role in maintaining transmission and helping reseed epidemics is expected to increase [19, 31]. In Esmeraldas, connectivity first increased with Colombia, consistent with the observation that the DENV serotypes circulating in Esmeraldas between 2010-2014 more closely matched those circulating in Colombia than the rest of Ecuador [17, 19, 29]. Lack of infrastructure often tracks with rapid urbanization, and often results in limited water access, garbage collection, and sewage, which are all arboviral disease risk factors.

The ecological conditions in regions where DENV has recently emerged may further increase DENV intensity. For instance, while in Guayas and Manabí, entomological surveys reveal low vector density during the dry season and larvae proliferation during the wet season [16], a household survey in Esmeraldas found a non-linear relationship between mosquito density and rainfall (unpublished data). Similarly, in the Amazon, there is not as strong a distinction between the wet and dry seasons, potentially enabling sustained arbovirus transmission year-round. The combined effects of Amazonian ecological conditions, urbanization, and population growth are evident in the Amazonian cities of Iquitos, Perú and Manaus, Brazil, where once *Aedes aegypti* and DENV were introduced and the cities were large enough, the conditions were primed for severe epidemics and sustained transmission [32]. In Iquitos, Perú which is mainly accessible by river barge, invasion of the *Aedes aegypti* species was facilitated by these river boats travelling from urban to peri-urban to rural areas [33]. For this reason, similar attention should be paid to the changes caused by the rapidly urbanizing Amazon provinces of Ecuador, which are increasingly affected by population growth, urbanization, and migration due to the expansion of the oil industry [34].

Our study has several limitations. We were unable to test whether changes in *Aedes* risk were temporally associated with changes in DENV transmission intensity, as data on ecological and mosquito density data were not available before 2013. Further, we were unable to determine changes in the serotype-specific intensity of DENV transmission due to lack of serotype-level case data. We were also unable to quantitatively measure the effect of changes in connectivity between regions and DENV transmission intensity due to lack of available data. Finally, due to low numbers of cases, we were unable to measure transmission changes for each separate province in the Amazon.

Here, we tracked the changes and drivers of DENV transmission across Ecuador using over 20 years of longitudinal case data, detailed entomological measurements, catalytic models of DENV FoI, and maximum entropy models for species distribution modeling. We show that coastal provinces with large, connected cities experiencing the highest and earliest increase in DENV transmission intensity. Areas that remained isolated like the northern coast and the Amazon regions more recently experienced a rise in DENV transmission and endemicity. However, all provinces with suitable ecological conditions were affected by the chikungunya and Zika pandemics and had hotspots for *Aedes* and arbovirus disease risk. Future local and national control strategies should consider the emergence and re-emergence of multiple arboviruses transmitted by *Aedes aegypti* as well as differences in the natural and built environments, including the rapid expansion of vector-borne diseases into areas that previously were minimally affected. As human movement, population growth, urbanization, and climate change intensify, more areas within diverse countries will be affected simultaneously. Thus, surveillance and vector control efforts should learn from the historical dynamics for dengue, Zika, chikungunya, recognizing the potential for novel emerging diseases to follow similar patterns and strategically develop control measures which consider interactions and shared risk factors for these diseases.

## Data Availability

The data sources and programs used for analyses are detailed in the Methods section. The code and data for all force of infection models will be made available on Zenodo at the time of publication.

## Acknowledgment

We thank the Ecuador Ministry of Health for providing the published dengue, Zika, and chikungunya prevalence data needed for this analysis.

## Funding

This research was supported by the Intramural Research Program of the National Institute of Allergy and Infectious Diseases (to LCK) and the National Institutes of Health Grant R01AI323721 (to JE and JC), and the Secretary of Higher Education, Science, Technology and Innovation (SENESCYT) Grant PIC-12-INH-002 (to PP and VC). LCK was supported in part by the Global Health Equity Scholars Program NIH Fogarty International Center training grant D43TW010540. The funders had no role in the study design, data collection and analysis, decision to publish, or preparation of the manuscript.

## Author contributions

Conceptualization: LCK, JC, VC, EQ, Formal analysis: LCK, EQ, VC, Data collection and curation: VC, EQ, PP, Methodology: LCK, VC, EQ, PA, PP, JC, Writing-original draft: LCK, SB, T-AH, Supervision/project administration: JC, JE, VC, PP, EQ, Writing-review & editing: all authors.

## Competing interests

The authors declare no conflict of interest.

## Notes

### Competing Interest Statement

The authors have declared no competing interest.

### Author Declarations

Human subjects data sets were from anonymized public health surveillance data collected by the Ecuadorian Ministry of Health as part of routine care. The Instituto Nacional de Estadistica y Censos (INEC) data used in this study is freely available from an institutional website (https://www.ecuadorencifras.gob.ec/camas-y-egresos-hospitalarios/) and is completely anonymous. According to the international good clinical and research practice and in accordance with Ecuadorian legislation on clinical investigation, ethical approval by an institutional review board was not required. The public surveillance data collected by the Ecuador Ministry of Health (Sistema Nacional de Vigilancia en Salud Publica Ecuador, ViEpi data) was made available for research purposes without confidential information (anonymous), under the "Organic Law of Transparency and Access to Public Information" (Article 4 of Ley Organica de Transparencia y Acceso a la Informacion Publica, LOTAIP, Law 24 of Article 81 of the Ecuadorian Constitution).

